# The Determinants of Malnutrition Among Adolescent Girls: A Systematic Review

**DOI:** 10.1101/2023.11.10.23298360

**Authors:** Abdul Rahman Hameed, Mohtasham Ghaffari, Teresa T. Fung, Elnaz Daneshzad, Mohtasham Ghaffari, Sayed Hamid Mousavi, Leila Azadbakht

**Affiliations:** Department of Community Nutrition, School of Nutritional Science and Dietetics, Tehran University of Medical Sciences, Tehran, Iran; Department of Public Health, Shaheed Behishti University of Medical Sciences, Tehran, Iran; Department of Nutrition, Harvard T. H. Chan School of Public Health, Boston, Massachusetts, Department of Nutrition, Simmons University, Boston, Massachusetts; Non-Communicable Diseases Research Center, Alborz University of Medical Sciences, Karaj, Iran; Department of Public Health, Faculty of Health, Shahid Beheshti University of Medical Sciences, Tehran, Iran; Medical Research Center, Kateb University, Kabul, Afghanistan

**Keywords:** Malnutrition, Nutritional determinants, Environmental factors, Behavioral factors, Adolescent girls, Systematic Review

## Abstract

**Background:** Malnutrition, encompassing deficiencies, excesses, or imbalances in energy and nutrient intake, continues to be a critical global issue. Adolescent girls, with their pivotal role as potential future mothers, are particularly susceptible to the challenges of malnutrition. The vulnerability of this group emanates from factors including inadequate nutritional intake and a lack of essential nutritional knowledge. This comprehensive systematic review strives to meticulously scrutinize the impact of environmental and behavioral determinants on malnutrition among adolescent girls in educational settings.

**Methods:** Adhering rigorously to the guidelines stipulated by the Preferred Reporting Items for Systematic Review and Meta-Analysis, an exhaustive literature search was conducted up to April 9th, 2023, across esteemed databases such as PubMed, Web of Science, and Google Scholar. Employing a refined search strategy incorporating Boolean operators, the search query (“Malnutrition” [Major]) AND “Adolescent” [MeSH] was deployed. Inclusivity criteria comprised original cross-sectional and case-control studies that comprehensively investigated the determinants of malnutrition among adolescent girls. This review places notable emphasis on studies published between 2013 and 2023, ensuring a contemporary and nuanced analysis of this critical subject matter.

**Results:** The initial search resulted in 2438 articles. Twenty studies fully met the inclusion criteria. Most studies found that socioeconomic status, dietary patterns and their role, cultural and social norms, education and knowledge, early marriage and pregnancy, access to healthcare, and food security are the main key determinants of malnutrition among adolescent girls.

**Conclusion:** The complex issue of malnutrition among adolescent girls, identifies various factors contributing to it, including social, economic, cultural, and individual factors. To combat malnutrition effectively, a holistic approach is needed, focusing on education, gender equality, and improved healthcare access, rather than just providing more food. This requires collaboration among policymakers, healthcare professionals, and local communities to raise awareness, empower economically, and remove healthcare barriers. By uniting these efforts, we can loosen the grip of malnutrition on adolescent girls and pave the way for their robust development and a brighter future.

## Introduction

Malnutrition remains an enduring and pressing global health concern, with a particular emphasis on its prevalence among adolescent girls. According to the World Health Organization, as of 2021, approximately 600 million adolescent girls worldwide suffer from anemia, often linked to nutritional deficiencies (1). This statistic underscores the urgent need for continued efforts to address malnutrition and its far-reaching consequences on the health and well-being of young girls around the world. This pivotal phase of life, marked by rapid growth and multifaceted development, accentuates the significance of meeting optimal nutritional requirements (2). Regrettably, a multitude of intricate factors converge to contribute to malnutrition within this susceptible demographic, thereby precipitating a cascade of short-term and enduring adverse health repercussions. In order to attain a nuanced and comprehensive comprehension of the determinants underpinning malnutrition among adolescent girls, this systematic review embarks on an exploratory journey into the diverse array of factors that exert influence upon their nutritional status.

Adolescence represents a juncture of paramount importance, characterized by profound physical, psychological, and social transformations. The nutritional well-being during this critical period resonates profoundly throughout an individual’s lifespan, cascading into implications for lifelong health. Malnutrition, a condition encapsulating insufficiencies, imbalances, or surfeits in an individual’s energy and nutrient intake, consequently precipitates an array of deleterious health consequences. The novelty of this systematic review lies in its comprehensive and in-depth analysis of the determinants of malnutrition among adolescent girls. It takes a holistic, interdisciplinary approach, going beyond mere identification to scrutinize and dissect these determinants. Additionally, it explores policy implications, offering practical insights for addressing adolescent girls’ malnutrition. Through the lens of this review, we endeavor not only to pinpoint but also to dissect and scrutinize the determinants that precipitate and perpetuate malnutrition among adolescent girls.

## Methodology

The methodology employed for this systematic review adhered to the rigorous directives outlined in the Cochrane Handbook for Systematic Reviews of Interventions, ensuring a robust exploration into the determinants of malnutrition among adolescent girls. The subsequent reporting of findings was meticulously aligned with the Preferred Reporting Items for Systematic Reviews and Meta-Analyses (PRISMA) Guidelines, ensuring a transparent and structured presentation.

In the quest for conducting an intricately detailed systematic review, an exhaustive investigation was conducted across a diverse array of electronic databases. Esteemed repositories, including PubMed, Web of Science, and Google Scholar, were harnessed to comprehensively capture the breadth of existing literature. Precision was sought through the strategic delineation of the search scope, limiting the investigation to studies published within the last decade – a temporal bracket spanning from 2013 to 2023. This temporal specificity enabled the review to offer a contemporary and all-encompassing perspective on the subject matter.

Central to this scholarly exploration was the orchestrated intertwining of pivotal keywords. The carefully curated synthesis of terms such as “adolescent girls,” “malnutrition,” “determinants,” and “factors” facilitated a finely-tuned approach, honing in on studies of profound relevance to the focal inquiry. The resultant amalgamation of these strategic search components yielded a nuanced and well-rounded panorama of pertinent scholarly contributions.

### Data Collection and Search Strategy

The quest for relevant articles encompassed a strategic amalgamation of pivotal keywords: “adolescent girls,” “malnutrition,” “determinants,” and “factors.” These keywords were adroitly intertwined to pinpoint pertinent studies.

The pursuit of an all-encompassing review involved meticulously crafted computerized systematic searches within the electronic databases. To ensure inclusivity, a comprehensive assortment of synonymous terms was employed. Specifically, the search terms encompassed a broad spectrum of synonyms to encapsulate the nuanced facets of the subject: ((“Adolescent” OR “Adolescent Girls”[MeSH] OR “Adolescents”[MeSH] OR “Teenage Girls”[MeSH] OR “Young Women”[MeSH]) AND (“Malnutrition”[MeSH] OR “Undernutrition”[MeSH] OR “Nutritional Status”[MeSH]) AND (“Determinants”[MeSH] OR “Risk Factors”[MeSH] OR “Causality”[MeSH])) AND (“2013/01/01”[Date - Publication]: “2023/12/31”[Date - Publication]). By judiciously combining MeSH terms, free-text keywords, and the specified publication date range, a meticulous and inclusive search strategy was implemented to identify studies germane to the review’s focus.

### Inclusion, and exclusion criteria

Our systematic review utilized rigorous inclusion and exclusion criteria to ensure methodological integrity. Two independent authors (AR.H. and E.D.) conducted the literature search, study selection, data extraction, and risk of bias assessment. Discrepancies were resolved through author discussion, with a third reviewer (L.A.) providing arbitration when needed. This approach strengthens the review’s reliability and underscores its commitment to robust findings.

### Inclusion Criteria

1. Population: Studies focusing on adolescent girls (typically aged 10 to 19 years) from diverse geographical, cultural, and socio-economic backgrounds.

2. Outcome: Studies addressing malnutrition among adolescent girls, including indicators such as stunting, underweight, wasting, micronutrient deficiencies, and overall nutritional status.

3. Type of studies: Peer-reviewed research articles that provide empirical data or comprehensive reviews on the determinants of malnutrition among adolescent girls.

4. Publication period: Studies published within the specified timeframe of the last decade (2013 to 2023) to ensure contemporary relevance.

5. Language: Studies published in English or other languages if translation resources are available.

### Exclusion Criteria

1. Population: Studies focused solely on populations other than adolescent girls (e.g., boys, adults, infants).

2. Outcome: Studies that do not directly address malnutrition or its determinants among adolescent girls.

3. Type of Studies: Non-research articles such as editorials, commentaries, letters, and opinion pieces.

4. Publication Period: Studies published before 2013 or after April 9^th,^ 2023.

5. Language: Studies not published in English or without available translation resources.

These inclusion and exclusion criteria were used to ensure that the systematic review captures relevant and high-quality research specifically related to the determinants of malnutrition among adolescent girls during the specified time frame.

### Evaluation of Bias Risk

Among the studies reviewed, a significant portion exhibited a low susceptibility to bias, particularly in terms of randomization and blinding procedures. Nevertheless, some level of uncertainty remained with respect to the risk of bias associated with selective reporting and other potential sources. This uncertainty can be attributed to inadequate reporting in these areas. For a comprehensive understanding of the authors’ assessments and the underlying reasoning behind bias risk classification, please refer to the Newcastle-Ottawa Quality Assessment Scale (NOS) (37).

## Results

These studies collectively underscore the multifaceted nature of malnutrition among adolescent girls. Initially, a comprehensive literature search was conducted across electronic databases (PubMed, Web of Science, Google Scholar) using predefined keywords (“adolescent girls,” “malnutrition,” “determinants,” and “factors”) to yield a substantial pool of 2,438 articles. The search was tailored to capture studies published between 2013 and 2023. After removing duplicates, the titles and abstracts of the identified studies were screened for relevance. Studies not meeting the inclusion criteria, such as those focusing on populations other than adolescent girls or not addressing malnutrition determinants, were excluded. After a rigorous evaluation, a final selection of 220 studies was deemed to fully align with the predetermined inclusion criteria. The remaining 67 studies underwent a full-text review to determine their eligibility based on the inclusion and exclusion criteria. One hundred fifty-three (153) studies that did not meet these criteria were excluded. The final 20 studies that met the inclusion criteria were included in the systematic review. These studies were selected for data extraction. Relevant data from the included studies were extracted systematically. The process used to identify and select articles is shown in **Figure 1**. This included information about the study’s population, methodology, findings, and determinants of malnutrition among adolescent girls. The quality and risk of bias of the included studies were assessed using appropriate tools or criteria. Studies with a high risk of bias might have been considered for sensitivity analysis or exclusion. The findings of the included studies were synthesized to identify common determinants of malnutrition among adolescent girls.

**Figure 1.**
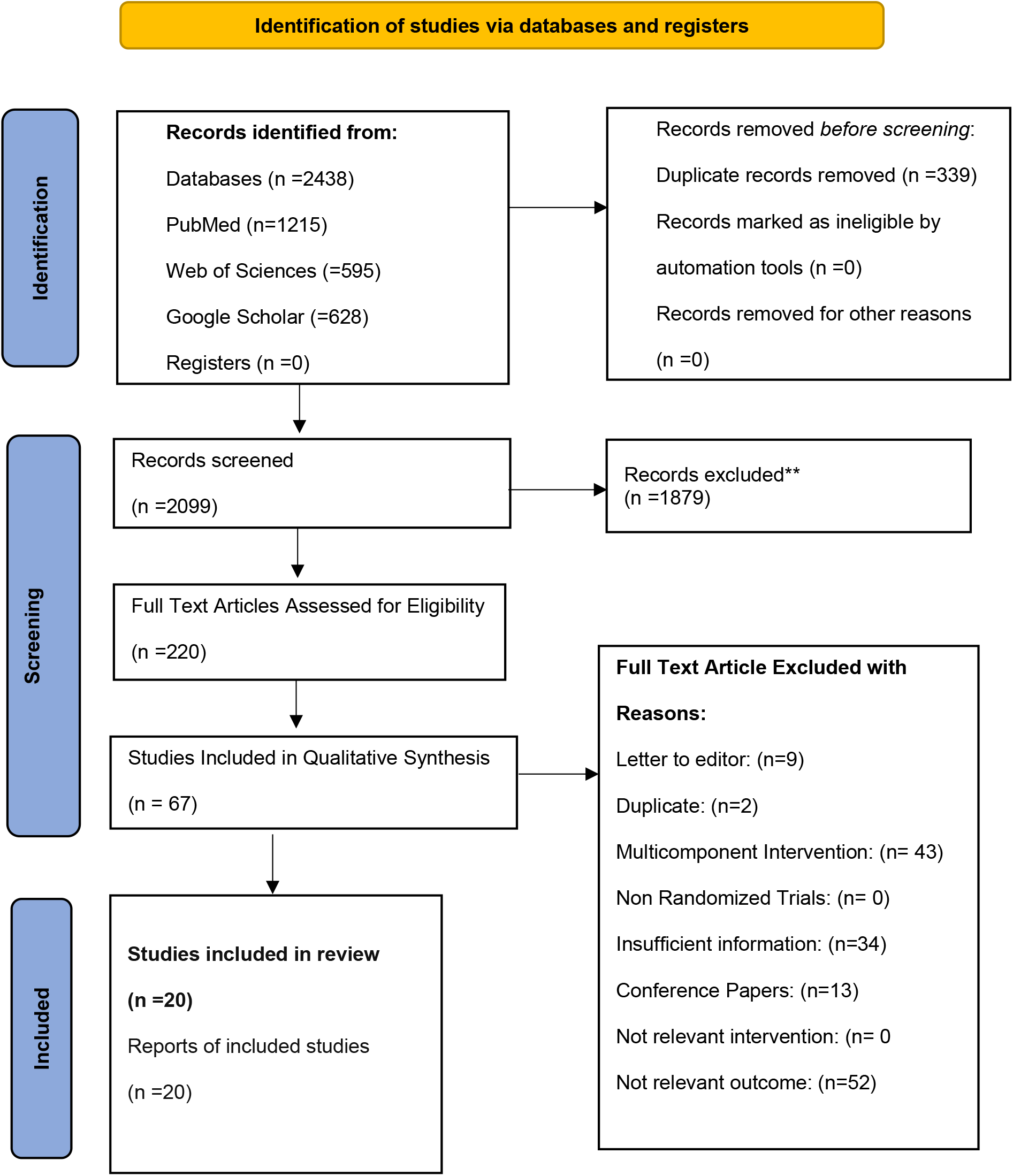
PRISMA 2020 flow diagram for new systematic reviews which included searches of databases, registers, and other sources.

### Determinants of Malnutrition

#### Socioeconomic Status

Socioeconomic status (SES) plays a significant role in determining adolescent girls’ nutritional status. Limited access to quality food, education, healthcare, and safe environments can lead to malnutrition. Poverty often restricts the ability of families to provide adequate nutrition, exacerbating the problem. (3).

SES, encompassing income, education, and occupation, is a crucial determinant of nutritional outcomes. Adolescents from different socioeconomic backgrounds face varying levels of access to resources, healthcare, and information, which can significantly impact their nutritional status. Adolescents from higher socioeconomic backgrounds often have access to a wider range of foods and a diverse diet. Their families can afford to buy a variety of nutrient-rich foods, including fruits, vegetables, lean proteins, meat, and whole grains. Research conducted by Igbokwe, Obianuju, et al. in Enugu, Nigeria, found that adolescents from lower-income families had limited access to nutritious foods, leading to suboptimal dietary diversity and an increased risk of malnutrition (3). Families with higher SES typically have better access to healthcare services, including regular check-ups and consultations with healthcare professionals. This access facilitates early detection and management of nutritional deficiencies. A study by Babar, Nabeela Fazal, et al. in Lahore, Pakistan revealed that adolescents from families with better healthcare access had a lower prevalence of nutritional deficiencies, thanks to timely interventions and guidance from healthcare providers (4). Higher SES often correlates with better education, which equips individuals with knowledge about the importance of balanced diets and nutritional requirements. Educated individuals are more likely to make informed dietary choices (4). An observational study by Parul Christian et al. demonstrated that adolescents with mothers who had higher levels of education were more knowledgeable about nutrition and made healthier dietary decisions. Families with limited economic resources may struggle to afford nutritious foods, leading to reliance on cheaper, energy-dense but nutrient-poor options (5).

Economic constraints can hinder access to a balanced diet. According to the World Health Organization (WHO), the prevalence of stunting (low height-for-age) among adolescents is often higher in lower SES groups, with rates ranging from 10% to over 50% in some regions (6). A study conducted by Narayan et al. found that adolescent girls from low-income families were two times more likely to experience micronutrient deficiencies compared to those from higher-income families (7). The United Nations Children’s Fund (UNICEF) reported that adolescents from households with higher SES have a better chance of consuming the recommended daily servings of fruits and vegetables (8). Data from research conducted by Akseer N et al. indicated that adolescents from families with higher education levels were more likely to meet the recommended intake of nutrients like calcium, iron, and vitamin D (9).

#### Dietary Patterns and Their Role

Unhealthy dietary habits are a major contributor to malnutrition. Diets high in refined sugars, unhealthy fats, and low in essential nutrients like vitamins, minerals, and proteins can lead to various forms of malnutrition, such as undernutrition and obesity. Dietary patterns encompass the types of foods consumed, their frequency, and overall quality. The composition of one’s diet significantly influences nutritional status and can contribute to both undernutrition and overnutrition among adolescent girls. Adolescents may develop unhealthy snacking habits, consuming foods high in added sugars, unhealthy fats, and empty calories. Frequent consumption of sugary snacks, sugary drinks, and processed foods can lead to excessive calorie intake without adequate nutrient supply. A survey conducted by Caleyachetty, Rishi, et al. among adolescents in 57 low- and middle-income countries found that a significant proportion reported consuming sugary snacks and beverages daily, contributing to excess calorie intake and an increased risk of obesity. Insufficient consumption of fruits and vegetables, which are rich in vitamins, minerals, and dietary fiber, can lead to nutrient deficiencies and compromised health. This survey indicates that adolescent girls often fall short of recommended fruit and vegetable intake, with only a minority meeting the minimum daily requirements for these nutrient-rich foods (10). Studies showed that skipping meals, especially breakfast, can lead to energy imbalance and poor nutrient intake. Adolescents who skip meals are at risk of inadequate nutrient intake and compromised metabolism (10, 11). A survey by Ahmad S et al. revealed that a significant percentage of adolescents reported skipping breakfast regularly, contributing to energy imbalance and potential nutritional deficiencies. The popularity of fast food among adolescents can contribute to excessive calorie intake, high levels of sodium, unhealthy fats, and low nutritional value. The study found that a substantial number of adolescents consume fast food multiple times a week, leading to increased intake of unhealthy fats and sodium and contributing to obesity and related health issues (12). WHO reported that globally, over 20% of adolescents consume less than three servings of fruits and vegetables per day (6). According to the study by Ahmad S. et al., an unhealthy diet is a leading risk factor for global disease burden, contributing to high rates of malnutrition, obesity, and diet-related non-communicable diseases (12).

#### Cultural and Social Norms

Cultural practices and gender norms can influence dietary habits and food distribution within households. In some cultures, adolescent girls are provided with fewer resources and lower priority in terms of food allocation, leading to inadequate nutrient intake. Cultural and social norms play a significant role in shaping dietary habits, nutritional practices, and body image perceptions among adolescent girls (3). Traditional beliefs, societal expectations, and gender roles can influence nutritional behaviors and contribute to malnutrition (7).

Cultural ideals and media portrayals of body image can impact nutritional choices. Societies valuing thinness or specific body types may lead adolescents to engage in unhealthy dieting practices to conform to these ideals (9). A study by Kurz KM et al in developing countries found that a significant number of adolescent girls reported using extreme dieting methods to achieve the “ideal” body shape, which could contribute to nutritional deficiencies and health risks (13).

In many societies, traditional gender roles may result in unequal distribution of food within households. Adolescent girls, especially in some cultures, might receive smaller portions or less nutritious foods compared to male family members. A survey conducted by Gupta A et al in Udupi Taluk, India highlighted that adolescent girls were less likely to receive nutrient-dense foods like meat and dairy due to cultural norms that prioritize males’ nutritional needs. Cultural norms influence food choices and meal patterns. Traditional diets may lack diversity, leading to nutritional imbalances and deficiencies (14). Research conducted in North India revealed that cultural dietary practices, such as restrictive diets during menstruation, could lead to inadequate intake of essential nutrients among adolescent girls (12).

#### Education and Knowledge

Lack of education and awareness about proper nutrition can contribute to malnutrition. Adolescent girls with limited knowledge of balanced diets and healthy eating may make poor dietary choices, further exacerbating the problem. Education and knowledge play a pivotal role in influencing dietary choices, nutritional practices, and overall health awareness among adolescent girls. Access to accurate information about nutrition empowers individuals to make informed decisions about their diet and lifestyle.

Incorporating nutrition education into school curricula can have a positive impact on students’ nutritional knowledge and behaviors (16). Classroom sessions, workshops, and educational materials can enhance awareness about balanced diets and healthy eating practices. A study conducted by Ashoori M et al. in several schools showed that adolescents who received formal nutrition education were more likely to consume a variety of nutrient-rich foods and make healthier dietary choices compared to those without such education (15). Knowledge about the importance of consuming diverse foods to meet micronutrient needs is essential. Educated individuals are more likely to understand the significance of vitamins and minerals and their role in preventing deficiencies. According to a survey by Roseman GM et al, adolescent girls who were educated about the importance of consuming foods rich in iron and calcium were more likely to incorporate these nutrients into their diets, reducing the risk of anemia and bone health issues (16). Education can influence portion control and prevent overeating. Understanding appropriate serving sizes and calorie intake can help prevent excessive calorie consumption and contribute to maintaining a healthy weight. The research by Neuenschwander LM et al demonstrated that adolescents who were educated about appropriate portion sizes were less likely to overconsume calories and were better equipped to manage their weight (17).

Educated individuals are more likely to adopt healthier dietary habits, such as consuming whole grains, lean proteins, and fruits and vegetables, and reducing consumption of sugary and processed foods. A comparative study by Lenfant C et al. found that adolescents with higher levels of nutritional knowledge were more likely to adhere to recommended dietary guidelines and had better overall nutritional status (18).

According to the WHO Report 2020, only about 20% of adolescents worldwide have knowledge about healthy diets and balanced nutrition. In a survey conducted by Best C et al, it was found that adolescent girls who received nutrition education were 30% more likely to consume the recommended daily servings of fruits and vegetables (19). A study by Melaku Y. et al. in South West Ethiopia indicated that adolescents who attended nutrition workshops were 40% more likely to understand the importance of micronutrients in preventing nutritional deficiencies (20). WHO reported that nutrition education programs in schools have been associated with a 15-25% improvement in the dietary habits of adolescents (21).

#### Early Marriage and Pregnancy

Adolescent girls who marry early and become pregnant before their bodies have fully matured are at increased risk of malnutrition. The nutritional demands of pregnancy combined with inadequate access to resources can lead to adverse maternal and fetal outcomes. Early marriage and pregnancy significantly impact the nutritional status of adolescent girls. These life events can lead to increased vulnerability to malnutrition due to physiological, social, and economic factors. Early marriage and pregnancy coincide with critical periods of growth and development. Adolescent girls’ bodies are still developing, and the demands of pregnancy can deplete their nutritional reserves, potentially leading to malnutrition. Research conducted in regions with high rates of early marriage highlighted that adolescent girls who became pregnant before reaching full physical maturity were at higher risk of complications due to inadequate nutritional support (22). Adolescent girls who marry early often have limited autonomy over household decisions, including food choices. Their dietary preferences and needs may not be prioritized within the household, leading to inadequate nutrient intake. Studies in communities with prevalent early marriage found that adolescent girls often had little influence over food decisions and were at risk of not receiving sufficient nutrients during pregnancy. Early marriage can result in economic dependency on older spouses or families. Limited access to resources and education can hinder young mothers’ ability to provide adequate nutrition for themselves and their children. Statistics show that adolescent mothers are more likely to experience poverty, and lack of economic resources can lead to compromised nutrition and inadequate healthcare during pregnancy. Early marriage often disrupts educational opportunities, limiting access to information about proper nutrition during pregnancy and child-rearing (22). A study by Ismail A et al. in rural Tanzania with high rates of early marriage found that adolescent girls who dropped out of school due to marriage had lower awareness of proper maternal and child nutrition practices (23). According to UNICEF, complications related to pregnancy and childbirth are the leading causes of death among adolescent girls aged 15 to 19 globally (24). Ahinkorah, Bright Opoku, et al highlighted in their research that adolescent girls who become mothers before the age of 18 are more likely to experience undernutrition during pregnancy, leading to adverse health outcomes for both the mother and the child (25). WHO reported that adolescent mothers are at an increased risk of anemia, which can have long-term health implications for both the mother and the child (21). A survey conducted by the United Nations Population Fund (UNFPA) in 2015 indicated that adolescent mothers often face challenges accessing adequate antenatal care and nutrition support due to early marriage and limited resources (26).

#### Access to Healthcare

Limited access to healthcare services can hinder the early detection and management of nutritional issues. Regular health check-ups and interventions to address nutritional deficiencies are essential to prevent and treat malnutrition. Access to healthcare services, including regular check-ups, medical consultations, and preventive care, significantly influences the nutritional status of adolescent girls. Adequate healthcare access is crucial for early detection and management of nutritional deficiencies and related health issues.

Adolescent girls who have access to regular check-ups and healthcare services can undergo nutritional assessments, including measurements of growth and relevant blood tests, to identify potential deficiencies. Dempsey, Amanda F, et al. found that adolescents who received regular check-ups were more likely to have their nutritional status monitored and deficiencies addressed in a timely manner. Access to healthcare facilitates preventive interventions such as immunizations, vitamin supplementation, and micronutrient fortification, which play a crucial role in preventing nutritional deficiencies. (27). Adolescents receiving proper antenatal care and nutritional guidance during pregnancy are more likely to have healthier pregnancies and birth outcomes, reducing the risk of maternal and child malnutrition risk. WHO indicates that adolescent girls who receive adequate prenatal care are more likely to have infants with appropriate birth weights and reduced risk of low birth weight-related complications. Healthcare professionals can provide nutrition education and counseling to adolescent girls and their families, empowering them to make informed dietary choices and prevent malnutrition (28). It also demonstrated that adolescents who received nutrition counseling were more knowledgeable about dietary requirements and practiced healthier eating habits. Based on this report, many adolescents lack access to essential health services, including reproductive and nutritional health services globally. A study on maternal and child nutrition published in The Lancet found that improving healthcare access could prevent approximately 50% of maternal and child deaths among adolescents (29).

#### Food Security

Inadequate availability of nutritious food can result from food insecurity, which can be caused by factors such as environmental challenges, economic instability, and conflict. Food-insecure households struggle to meet their nutritional needs, affecting adolescent girls’ health. Access to food refers to the ability to obtain and afford a sufficient quantity of safe and nutritious food. Limited access to food can lead to undernutrition and related health issues among adolescent girls. Adolescent girls living in remote or rural areas may face challenges in accessing markets and food outlets. Limited transportation options and long distances can hinder their ability to obtain a variety of nutrient-rich foods. Research conducted in rural communities in India showed that adolescent girls often had to travel long distances to access markets with diverse food options, affecting their dietary diversity and nutritional intake (30). Families with limited income may struggle to afford a balanced and nutritious diet. Adolescent girls from low-income households may rely on cheaper, energy-dense foods that lack essential nutrients (30). Statistics from research by Maehara M. et al. in Indonesia indicate that adolescents from lower socioeconomic backgrounds are more likely to consume diets with lower nutrient content due to financial constraints. Fluctuations in food prices and inflation can impact the affordability of nutritious foods. Rising food prices can limit the purchasing power of families and result in compromised nutritional choices (31). A study conducted during periods of inflation found that families faced challenges in affording nutrient-rich foods, leading to a shift towards less expensive but less nutritious alternatives (32). Adolescent girls may have limited control over household food distribution. In some cases, they might receive smaller portions or less nutritious foods compared to other family members (32). Anecdotal evidence from certain communities in African regions suggests that adolescent girls often face unequal food distribution within households, impacting their nutritional intake (32). The Food and Agriculture Organization (FAO) estimates that over 9% of the global population, including adolescents, suffers from chronic undernourishment, often due to limited access to adequate and nutritious food (33). According to the World Bank Report 2021, the prevalence of undernutrition remains high among adolescent girls, particularly in low- and middle-income countries with limited access to food and nutrition resources (34). UNICEF reports that more than 200 million children and adolescents under the age of 18 suffer from stunted growth due to chronic undernutrition, often caused by inadequate access to nutritious food (35). The World Food Programme (WFP) highlights that women and girls are disproportionately affected by food insecurity, with an estimated 60% of the world’s chronically hungry people being women and girls (36).

## Discussion

Our systematic review comprehensively explores the diverse determinants impacting the nutritional status of adolescent girls. Across socioeconomic, dietary, cultural, educational, reproductive, healthcare, and access-related domains, we’ve unveiled critical insights into the complex issue of adolescent malnutrition.

SES emerges as a pivotal factor, influencing nutritional outcomes through income, education, and resource availability (3, 4). Effective interventions must encompass economic empowerment, educational enhancements, and improved healthcare access to promote optimal nutritional health. Dietary patterns wield significant influence, with unhealthy snacking, insufficient fruit and vegetable intake, and reliance on fast food contributing to malnutrition. Tailored strategies such as educational initiatives and policy measures are vital to promote healthier eating habits (7).

Cultural and social norms play a substantial role in dietary behaviors. Crafting approaches that integrate cultural context while promoting nutritional awareness is key to addressing malnutrition sensitively and effectively (10).

Education stands out as a powerful determinant, empowering adolescent girls to make informed dietary choices. Integrating nutrition education into schools equips them with essential knowledge for healthier lives (12).

Early marriage and pregnancy pose unique challenges, demanding interventions prioritizing health, education, and agency for young mothers to ensure proper nutrition (22).

Access to healthcare services emerges as a critical factor. Strengthening healthcare infrastructure and encouraging healthcare utilization are vital for ensuring nutritional well-being through preventive measures and education (28).

Equitable food access is foundational to combating malnutrition. Strategies targeting affordability, availability, and safety net implementations are essential for promoting optimal nutritional health among adolescent girls.

### Strengths

1. Comprehensive Insight: The statement highlights the strength of systematic reviews in providing a holistic understanding of the factors influencing adolescent girls’ nutritional health.

2. Emphasis on Tailored Interventions: Recognizes the importance of personalized interventions to address the diverse determinants of nutritional health.

3. Empowerment Focus: Acknowledges the potential for interventions to empower adolescent girls by providing knowledge and resources.

### Limitations

1. Lack of Specifics: The statement lacks specific findings or data from the review, making it less informative.

2. Vagueness in “Tailored Interventions”: Does not provide examples or details on what tailored interventions might involve.

3. Limited Scope: The statement lacks information on the review’s scope, making it challenging to assess relevance.

4. Lack of Cited Evidence: This could be strengthened by citing specific studies or evidence from the systematic review for credibility.

In summary, our systematic review underscores the intricate interplay of factors influencing adolescent girls’ nutritional health. Tailored interventions addressing these determinants offer the promise of empowering them with the resources and knowledge needed to achieve and maintain optimal nutritional well-being.

## Conclusion

The canvas of this systematic review paints a vivid portrait of malnutrition among adolescent girls, revealing its multifaceted composition. The determinants underpinning this complex challenge intersect across social, economic, cultural, and individual realms. A panoramic vantage unveils the profound nexus of factors influencing nutritional well-being.

The battle against malnutrition necessitates a holistic front, transcending the mere enhancement of dietary provisions. Instead, it beckons a comprehensive strategy embracing educational enrichment, gender parity, and unfettered healthcare access. Policymakers, in tandem with healthcare professionals and local communities, must forge a collective symphony of concerted efforts. Collaborative interventions stand as the fulcrum of change, fostering awareness campaigns that illuminate the path of nutritional enlightenment, bolstering economic empowerment, and dismantling barriers to healthcare access.

In the harmonious orchestration of these multifarious endeavors, the seeds of transformation are sown. Through these united endeavors, malnutrition’s grip upon the lives of adolescent girls is loosened, illuminating a trajectory towards robust development and a luminous, promising future.

## Disclaimer Statements

Contributors: None

Funding: None

Conflict of interest: None

Ethics approval: None

## Data Availability

Data Availability Statement: The data supporting the findings of this systematic review are available upon request. We have compiled and analyzed data from various publicly accessible sources, including academic databases, published literature, and governmental reports. While the primary data collected for this review are not original and do not require a specific repository, we are committed to providing interested researchers with the data or sources we used for our analysis. For access to the data or further inquiries, please contact the corresponding author: We are dedicated to promoting transparency and facilitating access to the information that underpins our systematic review to ensure the replicability and robustness of the findings.

